# Amplicon-based nanopore minion sequencing of patients with COVID-19 omicron variant from India

**DOI:** 10.1101/2021.12.27.21268364

**Authors:** Somesh Kumar, Avinash Lomash, Mohammed Faruq, Oves Siddiqui, Suresh Kumar, Seema Kapoor, Prashanth Suravajhala, Sunil K Polipalli, SCOG_MAMC_ LNH

**Author notes:** Equal Contributors (Somesh Kumar, Avinash Lomash, Sunil K Polipalli).

## Abstract

SARS-CoV-2 infection has been playing havoc with emerging omicron variants of concern (VoC). Here, we report sequencing of the omicron variant in 13 patients from India using Oxford Nanopore Technology (ONT) Minion, wherein a rapid amplicon based sequence analysis was performed to assess and compare with existing 34 mutations in spike glycoprotein. We highlight and discuss the nature of these mutations that are unique and common to other populations. This is perhaps the first report on omicron variants from India using a long read sequencing chemistry.

## Introduction

Coronavirus Disease (COVID-19) is caused by severe acute respiratory syndrome coronavirus 2 (SARS-CoV-2) and has been an unfolding saga as the most discussed disease in recent times (www.who.int last accessed December 23, 2021; Zheng et al. 2020; Kaur et al., 2021). As of December 23, 2021, there have been 277,575,992 confirmed cases of COVID-19, including 5,395,365 deaths, reported to WHO. Although over 8,158,815,265 vaccine doses have been administered, with the emergence of delta and recent omicron variants, there have been several deaths all over the world. As re-infections are on rise, the scientific community has focused its attention to understand and gain evidence in lieu of wading immunity (Kai and Gratchen et al, 2021). As on December 23, 2021, over 87 countries have reported omicron variants ever since the first case was reported on December 1, 2021 (Bai et al. 2021). India has also seen 213 omicron cases with 4,79,000 deaths and over 29.8 million recovered so far (as on December 23, 2021). Early signs from previous reports indicate that omicron causes mild symptoms and the incidental findings would allow us to discern the modality behind emerging variants from India. Understanding the omicron variants would also help facilitate better prophylaxis on comorbidities with clinical presentations thereby distinguishing patterns associated with infection.

## Materials and Methods

### Patients’ samples

The nasopharyngeal swab samples were collected from 13 patients who have had a travel history of Africa/Middle-East. The patients were presented with mild symptoms within three days of onset of infection prior to hospitalization. Informed consent was judiciously taken before the sample was sequenced.

### Detection and sequencing of SARS-CoV-2 samples

The nucleic acid from these cases were subjected to Oxford Nanopore Technologies (ONT) long-read whole genome sequencing by preparing libraries using the midnight protocol of ARTIC classic approach (Catalog id SQL-RBK-110-96). The approach entails a series of short steps by sequencing and simultaneously mapping the reads to the reference genome besides analyzing the output data in real time (https://nanoporetech.com/covid-19; assessed December 14, 2021). The rapid 5-hour workflow, with 15-min fast library preparation was applied for backward tracing of the strains out of the lab, bringing genomic/molecular epidemiology analysis for discovery of mutations. Using the PCR tiling approach, the viral genome was amplified in overlapping sections across the full length of the genome with the Minion flow cell (FLO/106), following which base calling was performed; reads mapped and filtered to the Wuhan reference genome. The samples were labeled with in house registry identities and further named as LNHD1, LNHD2 and so on.

### Data pipeline and phylogenetic analysis

After the library preparation was done, careful execution of bar codes were supplied with the samples which were later run through MinKnow for approximately 3 hours so as to obtain fast5 files. The fast5 files were converted to fastq and a final annotation report was obtained through a series of steps. Commander was used to streamline the report as the workflow ensured all barcodes were run through quality check, amplicon coverage, genome coverage, variant calling and VCF file generation (https://genotypic.co.in/Commander; last accessed December 23, 2021). A 20x mean coverage of ca. 96.28% raw reads were mapped to SARS-CoV-2 reference genomes (accessions MN908947.3/NC_0145512), and a few sequences indicative of variable coverage (‘N’ or ‘X’). Using the initial set of these sequences, we have taken ‘‘good-sequence’’ alignment of all 13 omicron cases from India aligned to Wuhan reference, and a neighborhood joining tree was constructed using clustal omega with the sequences sorted vertically, thereby drawing a circular and unrooted tree (Sievers et al. 2011; Figure 1c/d). The tree was later visualized using interactive tree of life (iTOL; etunic and Bork et al. 2021). Finally, the GIASIA’s audacity instance was used to identify the mutations in spike glycoprotein and calculate the neighbor distance between related sequences, and datawrapper (datawrapper.de last accessed December 23, 2021) was used to plot the sequence data. All the 13 sequences were uploaded to the Global Initiative for Sharing All Influenza Data (GISAID) (Elbe and Buckland-Merrett, 2017) submission making data available for free access.

**Figure 1:**
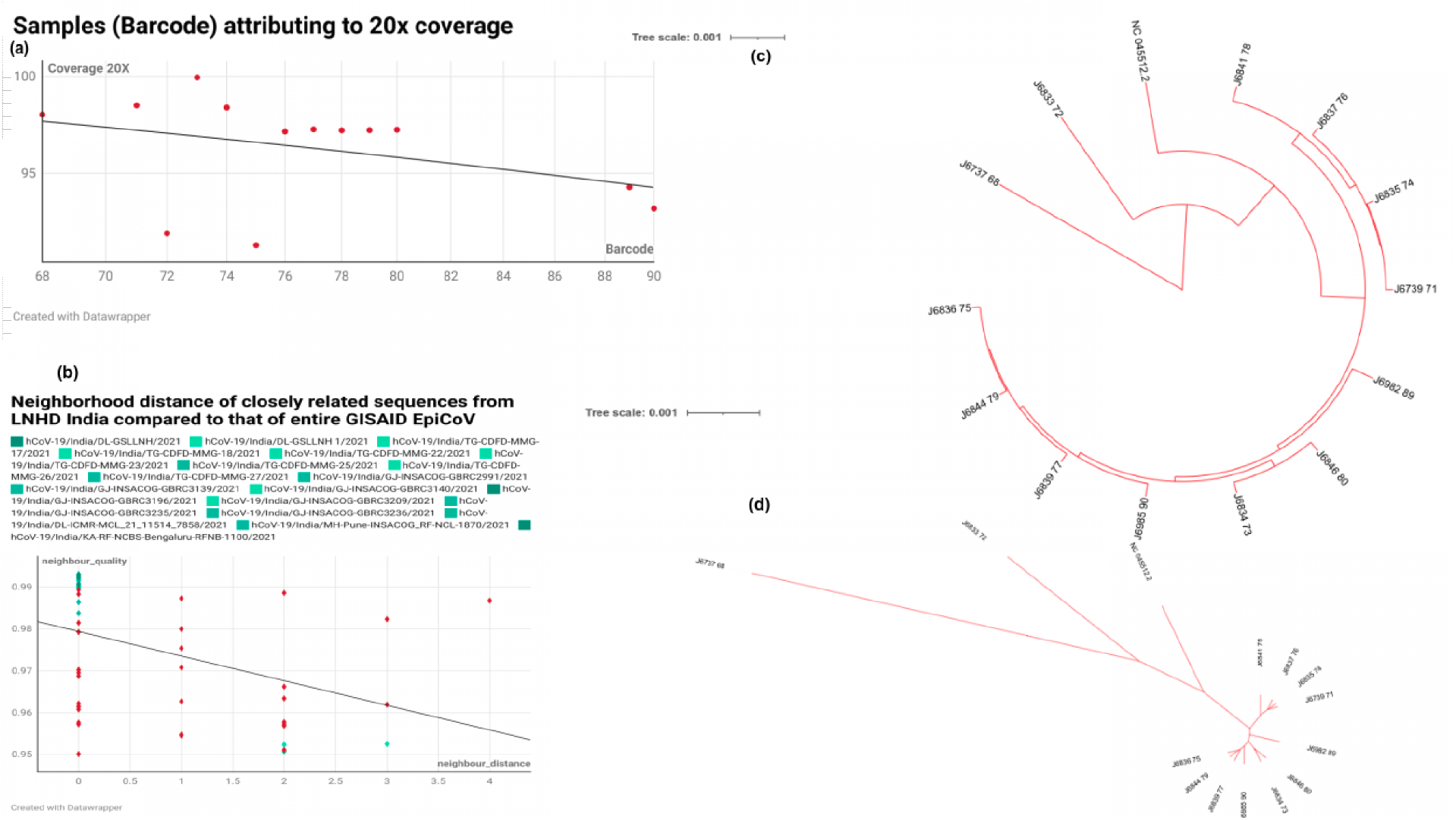
(a) Samples attributing to 20X coverage; (b) Neighborhood distance with quality check of closely related sequences (c)Circular phylogenetic tree of all 13 samples from India claded with Wuhan reference genome. (d) Unrooted tree showing a clear dissection of Wuhan from other lineages

## Results and Discussion

The ONT minion yielded a total read length attributing to 25 million reads on an average from all 13 samples, and the genomic GC content varied from ca. 40 to 43% covering 96.28% of the SARS-CoV-2 genome (20x) (Table 1; Figure 1a). To check the transmissibility associated with the number of mutations in the spike glycoprotein associated with receptor-binding domain (RBD), we compared the 44 common mutations from our samples with the recently emerging mutations of omicron (Figure 2). Our preliminary analysis indicate that the omicron variant subcladed with the dominant Delta variant and might have evolved rapidly from the multiple mutations in spike proteins further interacting with the angiotensin converting enzyme 2 (ACE2) receptor or Ab/IFN-gamma (Supplementary Tables 1-4; Figure 2).

**Table 1:**
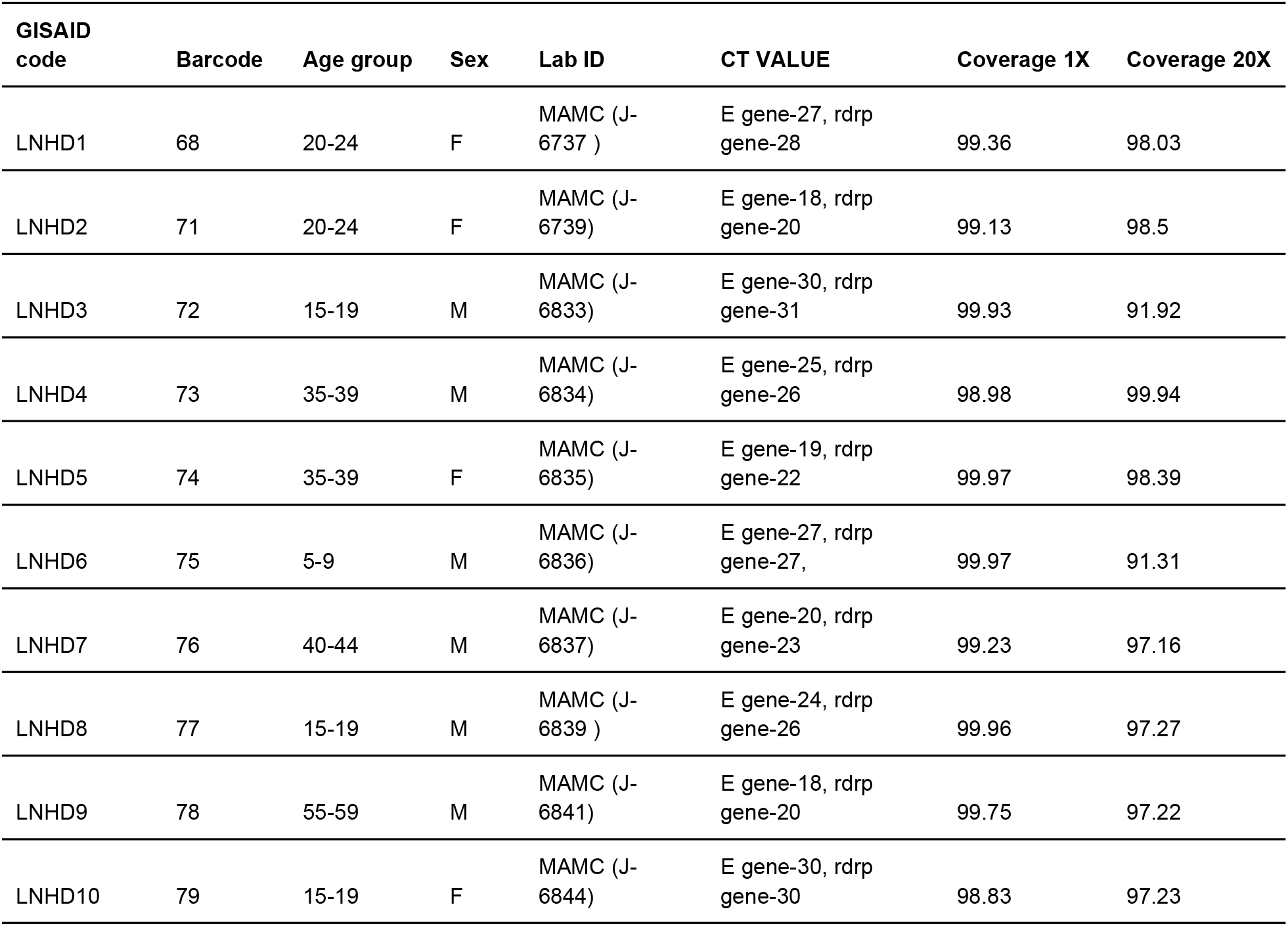
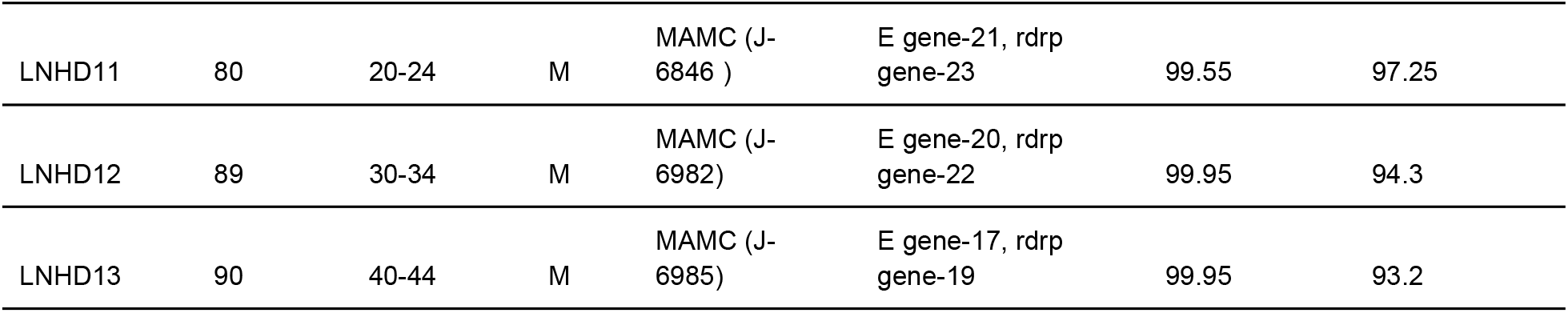
List of 13 samples with the coverage, CT values and age group in years/sex.

| GISAID code | Barcode | Age group | Sex | Lab ID | CT VALUE | Coverage 1X | Coverage 20X |
| --- | --- | --- | --- | --- | --- | --- | --- |
| LNHD1 | 68 | 20-24 | F | MAMC (J-6737 ) | E gene-27, rdrp gene-28 | 99.36 | 98.03 |
| LNHD2 | 71 | 20-24 | F | MAMC (J-6739) | E gene-18, rdrp gene-20 | 99.13 | 98.5 |
| LNHD3 | 72 | 15-19 | M | MAMC (J-6833) | E gene-30, rdrp gene-31 | 99.93 | 91.92 |
| LNHD4 | 73 | 35-39 | M | MAMC (J-6834) | E gene-25, rdrp gene-26 | 98.98 | 99.94 |
| LNHD5 | 74 | 35-39 | F | MAMC (J-6835) | E gene-19, rdrp gene-22 | 99.97 | 98.39 |
| LNHD6 | 75 | 5-9 | M | MAMC (J-6836) | E gene-27, rdrp gene-27, | 99.97 | 91.31 |
| LNHD7 | 76 | 40-44 | M | MAMC (J-6837) | E gene-20, rdrp gene-23 | 99.23 | 97.16 |
| LNHD8 | 77 | 15-19 | M | MAMC (J-6839 ) | E gene-24, rdrp gene-26 | 99.96 | 97.27 |
| LNHD9 | 78 | 55-59 | M | MAMC (J-6841) | E gene-18, rdrp gene-20 | 99.75 | 97.22 |
| LNHD10 | 79 | 15-19 | F | MAMC (J-6844) | E gene-30, rdrp gene-30 | 98.83 | 97.23 |
| LNHD11 | 80 | 20-24 | M | MAMC (J-6846 ) | E gene-21, rdp gene-23 | 99.55 | 97.25 |
| LNHD12 | 89 | 30-34 | M | MAMC (J-6982) | E gene-20, rdp gene-22 | 99.95 | 94.3 |
| LNHD13 | 90 | 40-44 | M | MAMC (J-6985) | E gene-17, rdp gene-19 | 99.95 | 93.2 |

**Figure 2:**
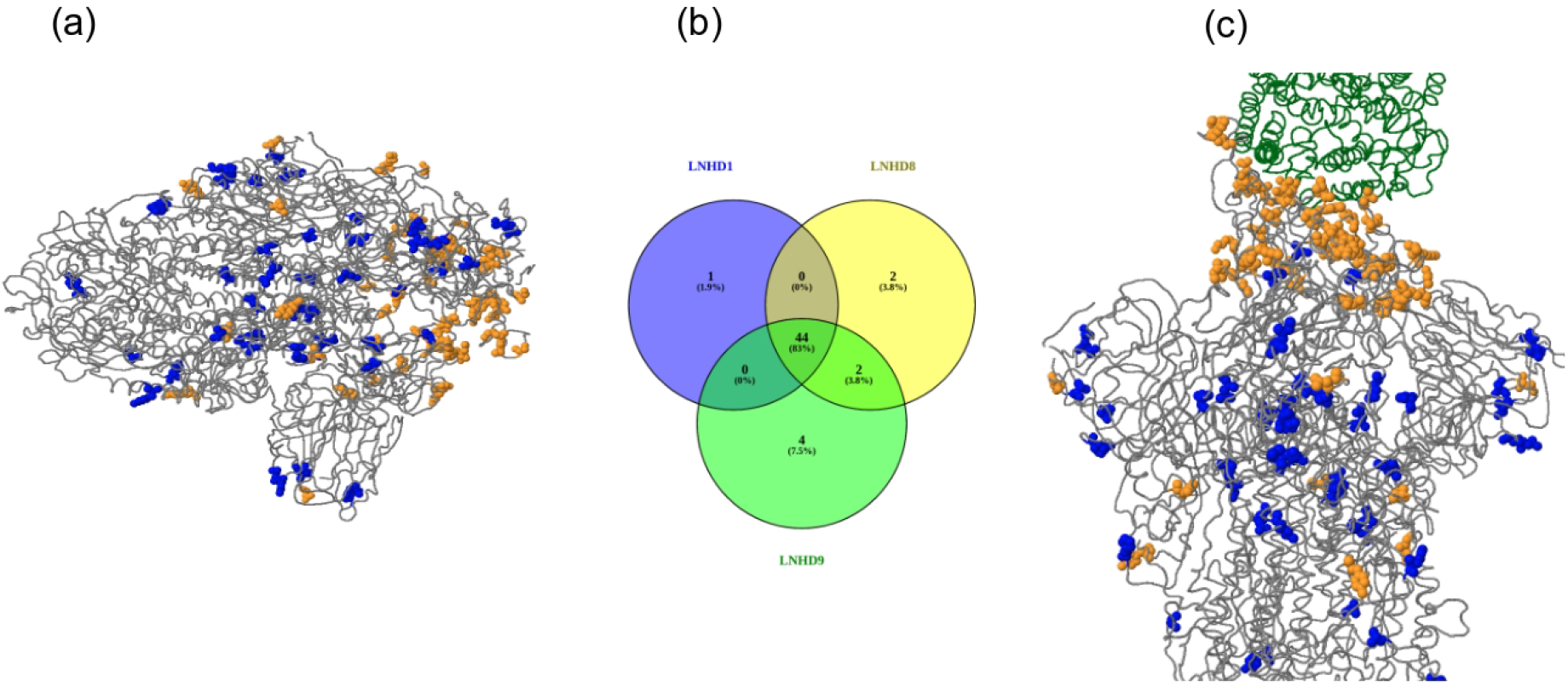
The spike mutations (#35) seen in Indian sub-population with the nearest residue if in loop/termini region (A67V, V70I(69), T95I, G142V, Y145H(143), N211I, L212I, G339D, R346K, S371L, S373P, S375F, K417N, N440K, G446S, S477N, T478K, E484A, Q493R, G496S, Q498R, N501Y, Y505H T547K, D614G, H655Y, N679K(674), P681H(674), A701V, N764K, D796Y, N856K, Q954H, N969K, L981F). (a) spike glycoprotein (PDB: 6acc, EM 3.6 Angstrom) with RBD in down conformation. (b) (b) Multivenn diagram of three samples LNHD1, LNHD8 and LNHD9 showing exclusively unique mutations in those samples and those common to all the LNHD series; (c) spike glycoprotein (PDB: 6acj, EM 4.2 Angstrom) in complex with host cell receptor ACE2 (green ribbon). *See supplementary Table 1 for details*

From the phylogenetic analysis, we observed that the Indian omicron variants were clustered together with a root emerging from OL815455, the variant that was first detected from Botswana. The iTOL containing the 13 sequenced samples and Wuhan reference yielded distinct clades both in unrooted and rooted circular tree, and the four samples that were claded separately perhaps indicate that they were among the first suspects of omicron in India (https://cov-lineages.org/lineages). With over 56 published omicron variants inGisaid.org from India (as on December 23, 2021), sequence comparison with Illumina short reads would allow us to understand the efficacy of third wave and inclination for possible herd immunity.

The neighborhood distance of closely related sequences from our LNHD series when compared to that of the entire GISAID EpiCoV database revealed that LNHD9 (with D155Y mutation) has a distinct clade with GBRC3290, a sample from Gujarat that was submitted recently. Both the samples with GR clade carry a combination of nucleocapsid, spike and NS3 mutations in addition to spike D614G (Sengupta et al. 2021). Taken together, we sought to ask whether or not any samples from our cohort have common and exclusively unique mutations. We obtained p.Thr614Ile, p.Thr1822Ile, p.Thr6098Ile, p.Asp155Tyr from LNHD9, p.Ala701Val and p.Val1887Ile from LNHD8 and p.Gly667Ser from LNHD1 (Figure 2c) with other samples having all common mutations reported (Supplementary Table 3). However, our preliminary observations indicate that none of these are known to be detrimental to the spike (Pires et al. 2014; data not shown). Mutations in the spike proteins of SARS-CoV-2 variants of concern (VoC) have also been compared to the parental SARS-CoV-2 isolate B.1 suggesting that the amino acid substitutions are already found in altered positions but with distinct substitutions. Finally, a large number of samples matching up to the sequence with neighbor distance of 0 and 2 (and a few with neighbor distance 4) showed that the mutational load or overburden could be attributed from delta or previous lineages (BA. 1; Supplementary Table 5)

The outbreak of COVID-19 caused by SARS-CoV-2 has swiftly spread worldwide with the probable origin of omicron still unclear. Interestingly, all the emerging case reports with no or mild upper respiratory tract symptoms are either with asymptomatic or oligosymptomatic transmission (Kupferschmidt K and Vogel G, 2021). The limitation of our study is that although the adopted ARCTIC protocol allowed the confirmation of SARS-CoV-2 infections, given the urgent need for rapid identification and traceability of omicron variants, an indefatigable checking the antibody titer against deleterious mutations and subsequent single cell genomic assay would have been a better proposition (Kannan et al. 2022). Furthermore, the difference in viral loads among samples will possibly affect the stability of average depth and genome-wide coverage with increase in whole-genome mapping. In summary, our study has demonstrated the utility of nanopore sequencing for SARS-CoV-2 genomes from clinical specimens. We firmly hope that prompt diagnosis and rapid whole-genome analysis would allow a decisive response to the SARS-CoV-2 outbreak that will bring disease control and prevention efforts.

## Data Availability

All omicron variant samples have been uploaded to GISAID.org with hCoV-19/India/un-LNHDXX/2021 series

https://www.gisaid.org/

## Acknowledgements

The authors gratefully acknowledge Government of Delhi and Institutional Ethics Committee of Maulana Azad Medical College, Delhi, India for granting the clearance of the study (F.1/IEC/MAMC/85/03/2021).

## Authors’ contributions

SK, AL, SKP, SCOG members performed library preparation and ran the ONT Minion., PS, SK, SKP and MF performed bioinformatics and genomic analysis., OS crosschecked the clinical spectrum of samples., SKu and SKa co-led the investigation and surveillance of the project., SK, SKP and PS wrote the draft and proofread the manuscript before all authors agreeing to the final version.

## Data availability

All omicron variant samples have been uploaded to GISAID.org with hCoV-19/India/un-LNHD**XX**/2021 series

SARS COV2 CONSORTIUM: Maulana Azad Medical College & Associated Lok Nayak Hospital (SCOG_MAMC_LNH)*

1. Dr Suresh Kumar, Medical Director, LNH and Head of Department of Medicine, Maulana Azad Medical College, New Delhi
2. Dr. Anurag Aggarwal, Director, CSIR, IGIB, New Delhi-
3. Dr. Sujeet Singh, Director, NCDC, New Delhi
4. Dr. Seema Kapoor, Director Professor, Department of Pediatrics, Maulana Azad Medical College, New Delhi.
5. Dr Sonal Saxena, Director Professor & Head of Department, Department of Microbiology, Maulana Azad Medical College, New Delhi.
6. Dr B.C.Koner, Director Professor and Head, Department of Biochemistry and Chairman, Medical research Unit Maulana Azad Medical College, New Delhi
7. Dr M.M. Singh, Director Professor, Dept of Community Medicine, Maulana Azad Medical College, New Delhi
8. Dr Vikas Manchanda, Professor, Department of Microbiology, Maulana Azad Medical College, New Delhi.
9. Dr Sandeep Garg, Professor, Department of Medicine, Maulana Azad Medical College, New Delhi
10. Dr Pragya Sharma, Professor, Dept of Community Medicine, Maulana Azad Medical College, New Delhi
11. Dr. Rajesh Pandey, Senior Scientist, CSIR IGIB, New Delhi
12. Dr. Mohamed Faruq, Principal Scientist, CSIR IGIB
13. Dr Sunil K Polipalli, Cytogeneticist, Genetic Lab, Department of Pediatrics, Lok Nayak Hospital, New Delhi.
14. Dr Oves Siddiqui, Associate Professor, Department of Microbiology, Maulana Azad Medical College, New Delhi.
15. Dr. Himanshu Chauhan, Additional Director, NCDC, New Delhi
16. Dr. Arvind Mohan, Medical Officer, Department of Medicine, Maulana Azad Medical College, New Delhi
17. Dr. Meenakshi Bothra, Assistant Professor, Department of Pediatrics, Maulana Azad Medical College, New Delhi.
18. Dr. Ranjana Mishra, Medical Officer, Thalassemia Screening programme, Lok Nayak Hospital. New Delhi.

## References

1. Bai Y, Du Z, Xu M, et al. International risk of SARS-CoV-2 omicron variant importations originating in South Africa. Preprint. medRxiv. 2021;2021.12.07.21267410. Published 2021 Dec 7. doi:10.1101/2021.12.07.21267410

2. Elbe, S., and Buckland-Merrett, G. (2017). Data, disease and diplomacy: GISAID’s innovative contribution to global health. Glob. Chall. 1, 33–46. GISAID: www.gisaid.org Last accessed: December 18, 2021.

3. Ivica Letunic, Peer Bork, Interactive Tree Of Life (iTOL) v5: an online tool for phylogenetic tree display and annotation, Nucleic Acids Research, Volume 49, Issue W1, 2 July 2021, Pages W293–W296, https://doi.org/10.1093/nar/gkab301

4. Kaur A, Chopra M, Bhushan M, Gupta S, Kumari P H, Sivagurunathan N, Shukla N, Rajagopal S, Bhalothia P, Sharma P, Naravula J, Suravajhala R, Gupta A, Abbasi BA, Goswami P, Singh H, Narang R, Polavarapu R, Medicherla KM, Valadi J, Kumar S A, Chaubey G, Singh KK, Bandapalli OR, Kavi Kishor PB, Suravajhala P. The Omic Insights on Unfolding Saga of COVID-19. Front Immunol. 2021 Oct 20;12:724914. doi: 10.3389/fimmu.2021.724914. PMID: 34745097; PMCID: PMC8564481.

5. Kannan SA, Austin N. Spratt, Kalicharan Sharma, Hitendra S. Chand, Siddappa N. Byrareddy, Kamal Singh, Omicron SARS-CoV-2 variant: Unique features and their impact on pre-existing antibodies, Journal of Autoimmunity. 126;2022 102779, ISSN 0896-8411, Kupferschmidt K, Vogel G. How bad is omicron? Some clues are emerging. Science. 2021 Dec 10;374(6573):1304–1305. doi: 10.1126/science.acx9782. Epub 2021 Dec 9. PMID: 34882443.

6. Pires DE, Ascher DB, Blundell TL. mCSM: predicting the effects of mutations in proteins using graph-based signatures. Bioinformatics. 2014;30(3):335–342. doi:10.1093/bioinformatics/btt691

7. Sengupta A, Hassan SS, Choudhury PP. Clade GR and clade GH isolates of SARS-CoV-2 in Asia show the highest amount of SNPs. Infect Genet Evol. 2021;89:104724. doi:10.1016/j.meegid.2021.104724

8. Sievers F., Wilm A., Dineen D., Gibson T.J., Karplus K., Li W., Lopez R., McWilliam H., Remmert M., Söding J., Thompson J.D. and Higgins D.G. (2011). Fast, scalable generation of high-quality protein multiple sequence alignments using Clustal Omega. Mol. Syst. Biol. 7:539

9. Zheng J. SARS-CoV-2: an Emerging Coronavirus that Causes a Global Threat. Int J Biol Sci. 2020;16(10):1678–1685. Published 2020 Mar 15. doi:10.7150/ijbs.45053

